# Analysis of Dental Tissues Density in Healthy Children Based on Radiological Data

**DOI:** 10.1101/2024.01.11.24301001

**Authors:** Reshetnikov Aleksei, Shaikhattarova Natalia, Mazurok Margarita, Kasatkina Nadezhda

## Abstract

**Introduction:** The information about the Hounsfield range values for healthy tooth tissues could become an additional tool in assessing dental health and could be used, among other data, for subsequent machine learning.

**Aim:** The aim of our study was to determine dental tissues density values in Hounsfield units.

**Materials and Methods:** The total selection included 36 healthy children (21 girls and 15 boys) with an age range of 6-10 to 11-15 years at the time of the study. The analysis of 320 teeth’s tissue density was carried out. The data were expressed as a mean and standard deviation. The significance was determined using the Student’s t-test. P-values less than 0.05 were considered significant.

**Results:** Analysis of the data showed that the tissues of healthy teeth in children have different density ranges. Enamel: 2954.69±223.77 HU - 2071.00±222.86 HU; dentin: 1899.23±145.94 HU - 1323.10±201.67 HU; pulp: 420.29±196.47 HU -183.63±97.59 HU. No gender differences concerning the density of dental tissues were reliably identified.

**Conclusion:** The evaluation of Hounsfield values for dental tissues can be used as an objective method for assessing their density. If the determined density values of the enamel, dentin, and pulp of the tooth do not correspond to the range of values for healthy tooth tissues, it may indicate a pathology.

**Key Messages:** Studying the range of healthy and diseased dental tissues using Hounsfield scores, as well as standardizing studies, can help clinicians improve screening accuracy and optimize follow-up of the effectiveness of preventive and therapeutic interventions in the future.

## Introduction

Nowadays, there are clinical, laboratory, and instrumental methods that allow not only to assess the initial condition of dental tissues but also to evaluate their changes during dental treatment.^[1,2]^ The monitoring of dental tissue condition is required in trauma, after transplantation, and during therapeutic procedures.^[3,4]^ It is especially important for children with metabolic diseases, genetic abnormalities, and children with special needs [5-7]. The use of innovative diagnostic methods provides dentists with new opportunities to assess dental health, especially in the early stages of pathological changes. Recently, cone-beam computed tomography (CBCT) has been widely used in dentistry.^[8]^ Unlike traditional orthopantomograms, CBCT allows the clinician to determine tissue density using Hounsfield units (HU).^[9,10]^ The information about tissue density for healthy teeth could become an additional tool in assessing dental health, their dental age estimation ^[11]^ and anatomy ^[12]^ and could be used, among other data, for subsequent machine learning.^[13]^ The results of earlier studies do not provide convincing data on the range of HU for healthy dental tissues in children of a certain age group.^[14,15]^ Our study is aimed at establishing Hounsfield values for dental tissues in children of a certain age group.

## Materials and Methods

The study protocol complied with the principles outlined in the Declaration of Helsinki of the World Health Organization and was approved by the Ethics Committees at Resto Dental Clinic Ltd (protocol number 07, December 22, 2020). Informed consent was obtained from the parents or legal guardians of all children under study. The study was conducted at Resto Dental Clinic Ltd, Izhevsk, Russia, from January 2021 to January 2023. The study included 35 children aged 6-10 and 11-15 of both sexes. The criteria for including children in the study were: 1) the presence of medical indications for CBCT (malocclusion and dental structural anomalies in the primary and permanent dentition, dental trauma, anomalies in dental position); 2) age 11-15 years; 3) consent to the study; and 4) the absence of genetic anomalies and concomitant diseases. Before CBCT, all subjects underwent a clinical study with a visual-tactile method. CBCT studies were performed using a Planmeca ProMax 3D tomograph (PLANMECA OY Asentajankatu 6, Helsinki, Finland) with scanning parameters of 88 kV, 5 mA, and 15 s. Only one expert clinician performed the measurements. PlanmecaRomexis 5.2.R 24.10.18 software was used to analyze the data obtained. The average value of dental tissue density was determined in an area of 1 mm2. Teeth of the upper and lower jaws that were selected for the study included: 1st permanent molars, permanent central incisors, 2nd primary molars, tooth germs of 2nd premolars, 2nd premolars, 2nd permanent molars, and tooth germs of 2nd permanent molars. Teeth enamel and dentin density were measured in HU on the incisor/occlusal surface (enamel 1, dentin 1) and proximal surface (enamel 2, dentin 2). The pulp density was measured in its central area (Fig. 1). The data were expressed as a mean and standard deviation.

The significance was determined using the Student’s t-test. P-values less than 0.05 were considered significant.

## Results

The total sample consisted of 36 healthy children (21 girls and 15 boys, (42%)) aged 6-10 and 11-15 years at the time of the study. The analysis of 320 teeth’s tissue density was carried out: 72 1st permanent molars, 72 permanent central incisors, 27 2nd primary molars, 40 tooth germs of 2nd premolars, 37 2nd premolars, 9 2nd permanent molars, and 63 tooth germs of 2nd permanent molars. Analysis of the data showed that the tissues of healthy teeth in children have different density ranges. Enamel: 2954.69±223.77 HU - 2071.00±222.86 HU; dentin: 1899.23±145.94 HU - 1323.10±201.67 HU; pulp: 420.29±196.47 HU -183.63±97.59 HU. No gender differences concerning the density of dental tissue were reliably identified. Statistical analysis did not reveal any significant relationships between Hounsfield values and demographic data (gender). Therefore, the density values of the tissues of the maxilla and mandible teeth were compared. Detailed data are presented in Table 1.

**Table 1.** Values in Hounsfield units of dental tissues density in healthy children.

|  | Enamel 1 | Enamel 2 | Dentin 1 | Dentin 2 |
| --- | --- | --- | --- | --- |
| HU, mean±standard deviation |  |  |  |  |
| 1st permanent molars |  |  |  |  |
| maxilla (n=36) | 2426.28±168.41 | 2358.81±219.60 | 1561.17±143.59 | 1584.11±137.17 |
| mandible (n=36) | 2414.53±194.85 | 2336.39±171.98 | 1537.50±150.25 | 1487.19±189.15 |
| * t-statistic | 0.2699 | 0.4756 | 0.6738 | 2.4543 |
| * t crit value | 2.0003 |  |  |  |
| permanentcentralincisors |  |  |  |  |
| maxilla (n=36) | 2954.69±223.77 | 2592.54±186.54 | 1796.40±163.39 | 1791.91±127.94 |
| mandible (n=36) | 2984.20±223.44 | 2552.37±186.85 | 1899.23±145.94 | 1871.69±98.81 |
| t-statistic | 0.5521 | 0.9001 | 2.7769 | 2.9202 |
| t crit value | 2.0003 |  |  |  |
| 2nd primarymolars |  |  |  |  |
| maxilla (n=16) | 2141.75±246.70 | 2228.53±160.24 | 1428.06±203.41 | 1413.21±145.79 |
| mandible (n=11) | 2227.09±115.66 | 2071.00±222.86 | 1323.10±201.67 | 1434.50±144.22 |
| t-statistic | 1.2045 | 1.9976 | 1.2867 | 0.3550 |
| t crit value | 2.0595 | 2.0639 | 2.0639 | 2.0739 |
| tooth germ of 2nd premolars |  |  |  |  |
| maxilla (n=21) | 2449.71±181.11 | 2509.62±221.56 | 1576.48±126.62 | 1649.71±128.85 |
| mandible (n=19) | 2583.68±134.75 | 2611.32±181.89 | 1666.42±138.10 | 1695.74±108.76 |
| t-statistic | 2.6698 | 1.5923 | 2.1394 | 1.2245 |
| t crit value | 2.0211 |  |  |  |
| 2nd premolars |  |  |  |  |
| maxilla (n=20) | 2220.58±190.65 | 2301.32±193.38 | 1417.30±119.57 | 1507.85±171.50 |
| mandible (n=17) | 2336.94±218.79 | 2348.18±103.87 | 1337.00±170.81 | 1375.18±126.17 |
| t-statistic | 1.7094 | 0.9365 | 1.6285 | 2.7042 |
| t crit value | 2.0211 |  |  |  |
| 2nd permanent molars |  |  |  |  |
| maxilla (n=4) | 2350.00±49.02 | 2403.50±101.93 | 1569.00±88.75 | 1523.25±91.31 |
| mandible (n=5) | 2293.40±131.28 | 2174.40±145.79 | 1443.00±70.81 | 1327.60±121.99 |
| t-statistic | 0.8897 | 2.7734 | 2.3111 | 2.7502 |
| t crit value | 2.3646 |  |  |  |
| tooth germ of 2nd permanent molars |  |  |  |  |
| maxilla (n=32) | 2359.03±169.39 | 2403.16±209.89 | 1527.03±121.39 | 1519.66±105.13 |
| mandible (n=31) | 2356.52±148.88 | 2499.97±178.51 | 1527.81±128.91 | 1554.37±120.88 |
| t-statistic | 0.0625 | 1.9741 | 0.0247 | 1.2031 |
| t crit value | 2.0423 |  |  |  |

\*The differences are significant at t-statistic > t critical value at p=0.05. \*\*t crit value - t critical value

The tissues (enamel and dentin) of permanent central incisors in the mandible and maxilla had the highest mean density value. The enamel and dentin density of the 2nd primary molars was significantly lower than 2nd permanent molars and tooth germ of 2nd permanent molars.

## Discussion

Nowadays, orthopantomograms, together with data from clinical, laboratory, and other instrumental studies, are used by clinicians to assess the condition of hard and soft tissues in the oral cavity in adults and children. However, the information content of orthopantomograms is limited to a 2D image, which may have plane distortions and does not allow accurate assessment of anatomical formations, such as teeth, in terms of their thickness, structure, and density. With the development of computed tomography and software, clinicians acquired an additional tool for determining the density of dental tissues.^[10]^ Previous studies of extracted teeth using micro-computed tomography showed an uneven distribution of enamel and dentin density in different areas of the tooth.^[13]^ Yavuz ^[14]^ confirmed this pattern in a mixed age population using CBCT in their study. However, the values of enamel and dentine density in their study were lower than the average values obtained during our study. One of the reasons justifying this difference may be the fact that our study included children of the same age group, which may justify further studies on the dental tissue density in a population of children and adults of certain age groups. The obtained density values for the tissues of tooth germs indicate that they correspond to the density of permanent teeth and exceed similar indicators of the density of primary tooth tissues. We believe that further research on the density range for healthy and pathologically altered dental tissues, as well as studies on standardization, can help clinicians improve the accuracy of screening and optimize subsequent monitoring of the effectiveness of preventive and therapeutic procedures in the future. This study is an attempt to establish a normal range of Hounsfield units for healthy maxillary and mandibular dental tissues in children of a certain age group. The data obtained revealed the density values for enamel, dentin, and pulp for primary and permanent teeth, as well as the germs of primary teeth. Differences in the density of specific teeth were also revealed; in particular, it was found that the enamel of the incisors has the highest density, significantly exceeding the density of the molars. Further research on the density of dental tissues in normal and pathological conditions seems promising, in particular for machine learning.

A limitation of our study is that measurements were carried out by only one expert clinician, which eliminates an assessment of interobserver variability. The study was conducted on a population of children in the same age group. In addition, not all maxillary and mandibular teeth were included in the study. The present study shows Hounsfield values of dental structures associated only with a particularly used CBCT machine. Further studies on a larger population, as well as on other age groups of patients, may be useful to improve the information content of the data.

## Conclusions

Evaluating Hounsfield values of dental tissues can be used as an objective method for assessing their density. Revealing density values of the enamel, dentin, and pulp of the tooth that do not correspond to the range of values of healthy tooth tissues may indicate their pathology.

## Data Availability

All data produced in the present work are contained in the manuscript

## Contribution Details (to be ticked marked as applicable)

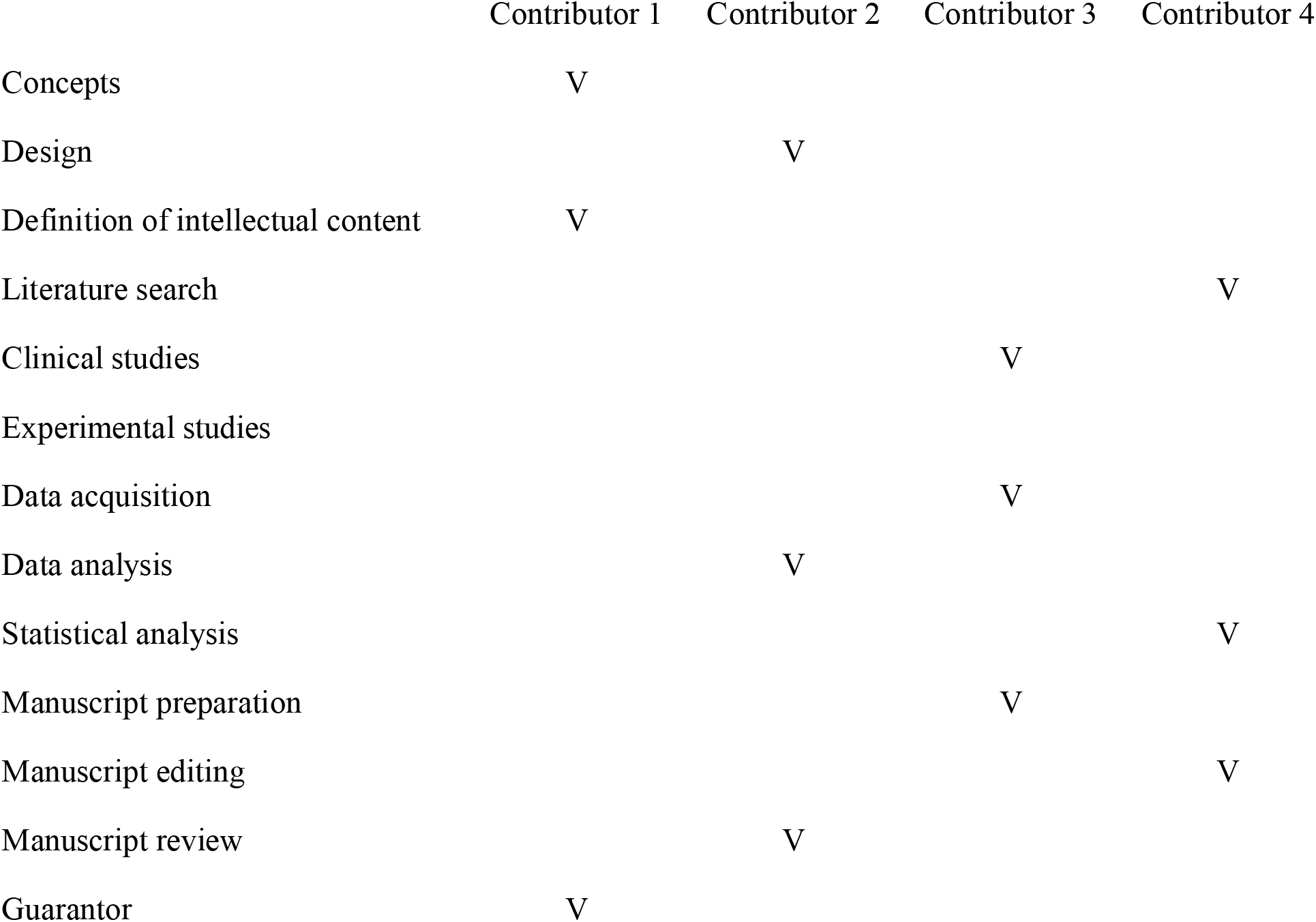

## Acknowledgement

The authors acknowledge and thank the Professor Aleksandr Urakov (from Izhevsk State Medical Academy) for thanks for all the support.

